# Persistence of training-induced visual improvements after occipital stroke

**DOI:** 10.1101/2024.10.24.24316036

**Authors:** Hanna E. Willis, Berkeley Farenthold, Rebecca S. Millington-Truby, Rebecca Willis, Lucy Starling, Matthew Cavanaugh, Marco Tamietto, Krystel Huxlin, Holly Bridge

**Affiliations:** Wellcome Centre for Integrative Neuroimaging, Nuffield Department of Clinical Neuroscience, University of Oxford, Oxford, United Kingdom, OX3 9DU; Flaum Eye Institute and Center for Visual Science, University of Rochester, Rochester, NY 14642, USA; Department of Psychology, University of Torino, 10123 Torino, Italy; Department of Medical and Clinical Psychology, Tilburg University, The Netherlands

**Keywords:** rehabilitation, visual training, stroke, restitution, hemianopia

## Abstract

Damage to the primary visual cortex causes homonymous visual impairments that appear to benefit from visual discrimination training. However, whether improvements persist without continued training remains to be determined and was the focus of the present study. After a baseline assessment visit, 20 participants trained twice daily in their blind-field for a minimum of six months (median=155 sessions), using a motion discrimination and integration task. At the end of training, a return study visit was used to assess recovery. Three months later, 14 of the participants returned for a third study visit to assess persistence of recovery. At each study visit, motion discrimination and integration thresholds, Humphrey visual fields, and structural MRI scans were collected. Immediately after training, all but four participants showed improvements in the trained discrimination task, and shrinkage of the perimetrically-defined visual defect. While these gains were sustained in seven out of eleven participants who improved with training, four participants lost their improvement in motion discrimination thresholds at the follow-up visit. Persistence of recovery was not related to age, time since lesion, number of training sessions performed, proportion of V1 damaged, deficit size, or optic tract degeneration measured from structural MRI scans. The present findings underscore the potential of extended visual training to induce long-term improvements in stroke-induced vision loss. However, they also highlight the need for further investigations to better understand the mechanisms driving recovery, its persistence post-training, and especially heterogeneity among participants.

## Introduction

Damage to V1 or its immediate afferent tracts, commonly caused by stroke, leads to loss of vision in the contralateral visual hemifield (Zhang *et al*., 2006). Every year, 12 million people in the world suffer from stroke, approximately one in four individuals in their lifetime (Feigin *et al*., 2022). Of these, an estimated 30-60% develop visual field deficits (Gray *et al*., 1989) which lead to impaired quality of life (Papageorgiou *et al*., 2007; Gall *et al*., 2009; Gall, Franke and Sabel, 2010). Clinical care is highly variable, and there are currently no widely used treatments for visual deficits post-stroke (see Willis and Cavanaugh, 2023 for recent review).

Despite this, over several decades, multiple studies have shown that visual training within the blind field can improve vision in the affected region in adult-onset V1 damage. Starting in the 1960s, researchers showed that non-human primates with V1 lesions were able to learn to detect and saccade to a point of light in their blind field (Cowey and Weiskrantz, 1963; Weiskrantz and Cowey, 1970; Mohler and Wurtz, 1977). These improvements occurred gradually with repeated testing, were location specific, and did not occur spontaneously (Weiskrantz and Cowey, 1970). Improvements in vision have been found for a range of visual stimuli, including flicker sensitivity (Raninen *et al*., 2007), target localisation (Chokron *et al*., 2008), orientation discrimination (Das, Tadin and Huxlin, 2014; Yang *et al*., 2024), letter identification (Raninen *et al*., 2007; Chokron *et al*., 2008), direction discrimination (Cavanaugh et al., 2015, 2019; Elshout et al, 2016; Willis et al., 2024), direction integration (Huxlin *et al*., 2009; Das, Tadin and Huxlin, 2014; Willis *et al*., 2024) and contrast sensitivity (Sahraie *et al*., 2006, 2010; Das, Tadin and Huxlin, 2014; Ajina *et al*., 2021; Yang *et al*., 2024). Moreover, studies have found that improvements can generalise to untrained tasks, with training on coarse direction discrimination improving contrast sensitivity (Das, Tadin and Huxlin, 2014) and fine direction discrimination (Cavanaugh *et al*., 2015). Additionally, training also reduces the size of the visual field deficit, as measured by clinical perimetry (Sahraie et al., 2006; Chokron et al., 2008; Huxlin et al., 2009; Elshout et al, 2016; Cavanaugh and Huxlin, 2017; Halbertsma et al., 2020; Willis et al., 2024).

These results are encouraging, suggesting that behavioural training may represent a promising intervention for stroke-induced vision loss. Moreover, in healthy controls, trained improvements in motion discrimination ability are retained for days (Sundareswaran, & Vaina, 1994) and even months after learning (Ball and Sekuler, 1987; Rokem and Silver, 2013). However, we currently know too little about the properties of recovered vision in stroke survivors with visual field deficits, including its longevity after the end of training and factors that may impact its persistence.

In non-human primates, V1 damage is indeed known to initiate a process of trans-synaptic retrograde degeneration across the early visual system - a degeneration of neurons unaffected by the initial insult but that degenerate as a result of losing their synaptic target. In this case, damage to V1 leads to gradual loss of LGN neurons (Van Buren, 1963; Cowey and Stoerig, 1989; Hendrickson *et al*., 2015; Atapour *et al*., 2017; Atapour, Worthy and Rosa, 2021). Fibres in the optic tract projecting to these degenerated regions of the LGN also begin to degenerate: studies in non-human primates and humans with V1 lesions found evidence of trans-synaptic retrograde degeneration of the ipsilesional optic tract (OT) (Bridge *et al*., 2011; Cowey, Alexander and Stoerig, 2011; Millington *et al*., 2014; Fahrenthold *et al*., 2021; Kim *et al*., 2022) and of the ganglion cell layer complex in the retina (Jindahra, Petrie and Plant, 2009, 2012; Yamashita *et al*., 2016; Schneider *et al*., 2019). Moreover, ipsilesional OT degeneration appears to correlate with lesion size and time since lesion (Cowey, Stoerig and Williams, 1999; Cowey, Alexander and Stoerig, 2011), and with early visual cortex activity for blind field stimuli, which predicts further ipsilesional retinal ganglion cell complex thinning six months later (Schneider *et al*., 2019). These results suggest that retrograde degeneration increases with time since damage – a progression that would be predicted to negatively impact blind field visual processing and prospects of benefiting from visual restorative interventions that stimulate the blind-field. Confirming this prediction, recent work showed that the level of OT degeneration measured prior to visual training predicts the amount of perimetric-based recovery attained (Fahrenthold et al., 2021). However, it is unknown whether the extent of trans-synaptic retrograde degeneration of the optic tracts relates to persistence of training-induced recovery.

Here, we performed a longitudinal training study in which occipital stroke survivors were trained on a motion discrimination task. They then stopped training for several months before repeat assessments of visual functioning and structure. This design allowed us to measure both the impact of a training intervention, as well as the longevity of training-induced visual changes. We then determined what, if any, biomarkers, or demographics might predict persistence of improvements.

## Methods

### Participants

Twenty-four stroke survivors with damage to V1 were recruited to take part in a visual rehabilitation study (Table 1, Figure 1, median age=49; range=24-71, 6 female). Pre-training magnetic resonance spectroscopy and behavioural data from these participants were analysed elsewhere (Willis *et al*., 2023). In addition, diffusion-weighted imaging data between the pre- and post-training visits in the same cohort were also recently published (Willis *et al*., 2024).

**Table 1.**
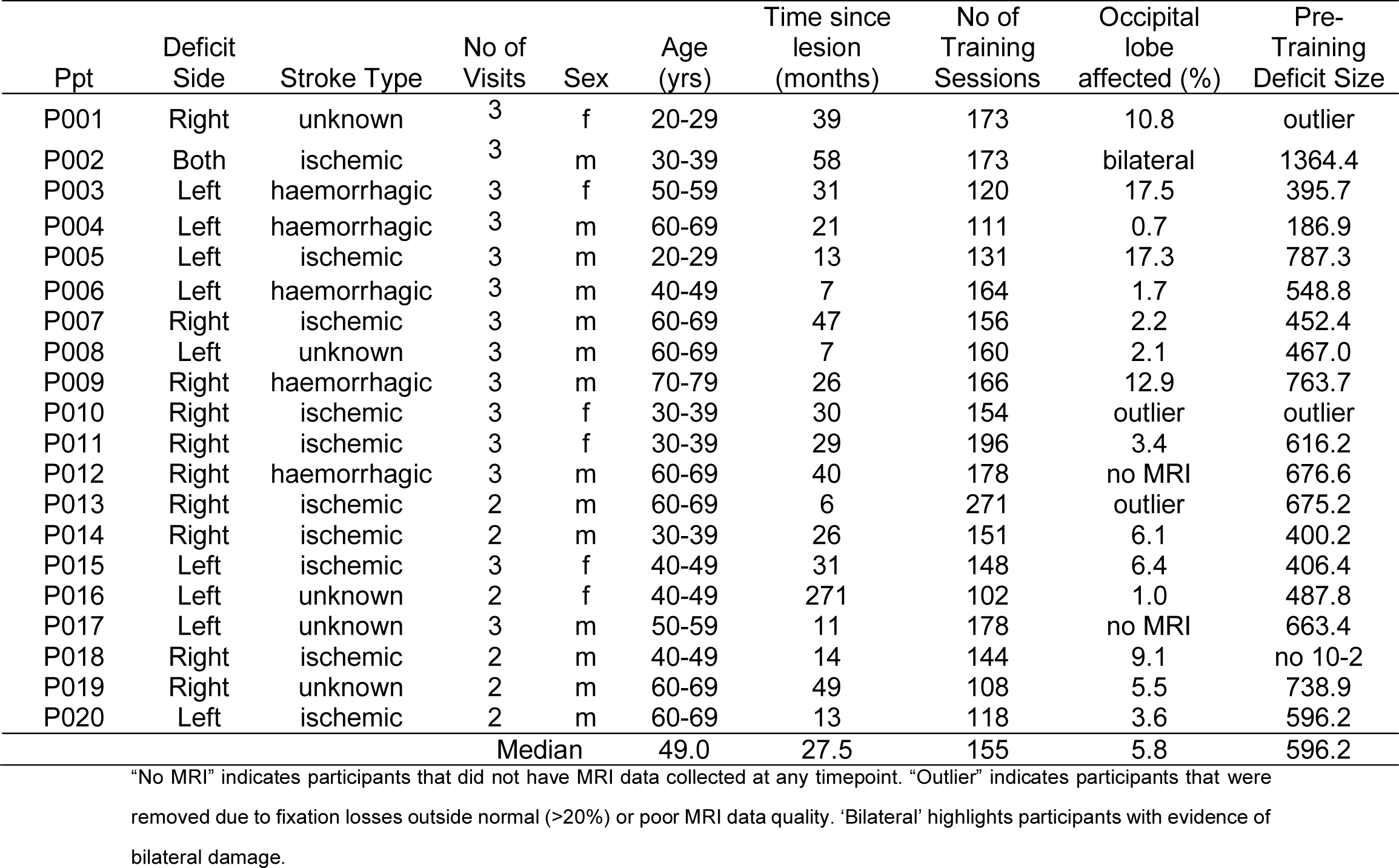
Participant demographics.

**Figure 1.**
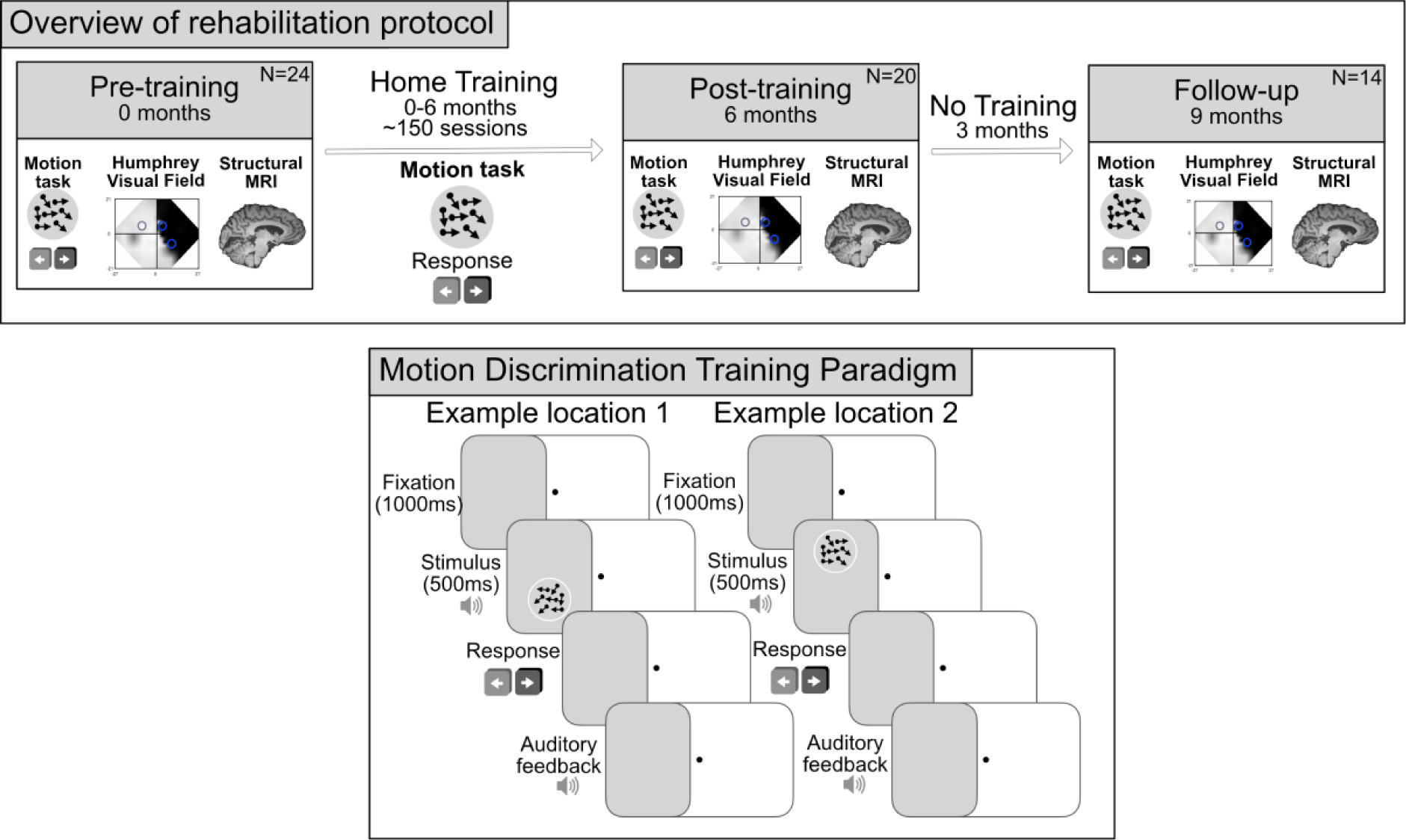
Overview of study design. (A) Study design involving three research visits; baseline (pre-training), after at least six months (post-training) and after a further three months (follow up). Participants completed six months of visual training at two locations in the blind field between the pre- and post-training timepoints. (B) Motion discrimination training paradigm: participants fixated centrally while motion stimuli were presented at a training location within the blind field. Participants were asked to discriminate the global direction of movement (leftward or rightward) of the dots in the stimulus after each presentation. Presentation of stimuli was accompanied by an auditory tone and auditory feedback signalled correctness of the response. This was repeated 300 times for each blind field locations.

Twenty of these participants completed at least 6 months of visual training in their blind field (minimum 100 sessions total) and were included in the present analyses. MRI data were collected in eighteen of these 20 participants, as 2 participants (P012, P017) were unable to perform the scan session.

Fourteen of the 20 participants then returned to the laboratory 3 months after completing training to determine whether training-induced improvements persisted across a period without training. Twelve of these participants also had MRI data collected at the follow-up visit, although the 2 participants (P012, P017) who were not scanned pre-training were also not scanned at other timepoints.

Importantly, participants that returned for the follow-up visit were not selected based on training progress or data quality; they simply finished training first and were able to return for another visit within the duration of the study.

### Inclusion criteria

Participants were healthy, English-speaking adults with stroke-induced damage to V1. Damage occurred in adulthood, resulting in homonymous visual field deficits (including hemianopia/partial hemianopia/quadrantanopia/scotoma). All participants were at least six months post stroke (i.e., in the “chronic” phase post-stroke). They did not have any diagnosed cognitive or psychiatric disorders, including executive or attentional deficits, or history of eye disease or impairment other than visual field deficits post-stroke, including all forms of visuospatial neglect. Lack of neglect was determined via telephone interview and medical records. There were no concerns about neglect for any participants. Participants did not undergo any other visual rehabilitation for the duration of the study.

### Ethics

Ethical approval was given for this study by the local ethics committee (R60132/RE001). Before taking part, participants read an information sheet and gave informed consent. The study was conducted in accordance with the ethical guidelines of the Declaration of Helsinki. Participants were made aware that visual training was to inform research and was not an established treatment and may therefore not guarantee improvement.

### Study design

Participants visited the Wellcome Centre for Integrative Neuroimaging, University of Oxford for a pre-training (N=24), post-training (N=20) and follow up (N=14) visit (see Figure 1 for schematic). Between pre- and post-training visits, participants were asked to perform two sessions of visual training per day (∼40minutes; 300 trials per location), at least 5 days per week for at least six months (Willis *et al*., 2024). On average, they completed 155 sessions (range=102-271), with a minimum requirement set at 100 sessions. Four participants did not complete the required number of sessions and were not included in this analysis. To ensure participants completed the minimum amount of training sessions, some continued training for longer than 6 months. For the post-training visit, participants returned on average eight months after their initial baseline visit *(median±IQR=8±3 months; range=6-14 months).* The amount of time elapsed between the pre- and post-training visits was affected by the individual’s rate of improvement, family or work commitments, scanner availability, and the participant’s ability to travel back to Oxford. This study was conducted across COVID-pandemic lockdowns and therefore, participants were not always able to return to lab at the appointed time. For the follow-up visit, participants returned approximately three months *(median±IQR=92.5±9.5 days; range=84-118 days)* after their post-training visit, with no training performed during this period. The study and planned analyses were registered on clinicaltrials.gov before the start of data collection (NCT04878861).

## Visual Training

### Software

Participants trained on a psychophysical global motion discrimination task using lab-issued chin-forehead rests, positioned 42cm away from their own laptop or external display screen. The training program was designed in MATLAB (MathWorks) using the Psychophysics Toolbox (Brainard, 1997; Pelli, 1997) and was identical to that used in previously published studies (e.g., Huxlin *et al*., 2009; Cavanaugh and Huxlin, 2017; Saionz *et al*., 2020; Willis et al, 2024). Software was customised for each participant’s computer or monitor specifications (including dimensions, resolution, and refresh rate). To ensure computer specifications were accurate, at the start of each training session, the programme presented a calibration square of a known size and participants were asked to measure the size of the box. At the pre-training visit, researchers ensured that participants learned how to use the training program appropriately. Participants were also given instructions detailing how to set the training up at home to ensure consistency. Online meetings were organised throughout home training to provide technical support or advice on setup, as needed. Participants were encouraged to train when they were most awake, and the exact time was recorded in the daily logs produced by the training programme.

### Training task

Task and stimulus parameters were identical to previous studies (Huxlin *et al*., 2009; Cavanaugh and Huxlin, 2017; Saionz *et al*., 2020; Willis *et al*., 2024). Random dot stimuli consisted of black dots on a mid-grey background (dot speed 10 degree/s, dot lifetime 250ms, stimulus duration 500ms; aperture 5 degrees diameter). Participants were asked to maintain fixation centrally on a dot while making coarse left-right discriminations of the global direction of motion in the random dot stimuli. The difficulty of the task was modulated using a 3:1 staircase procedure, which adjusted the direction range of the dots. After three correct responses, the direction range increased from 0 to 355° in 40° steps; after one incorrect response, it was decreased by 40°. An auditory tone accompanied stimulus presentation, and a distinct set of tones signalled correct and incorrect responses for each trial. After each home-training session, the software automatically generated a log file detailing trial-by-trial information and performance. These logs were emailed to the laboratory each week so that progress could be tracked. Direction range thresholds were calculated by fitting a Weibull function to the data with a criterion of 72% correct. Each threshold was then normalised to the maximum range of dot directions (360°) to generate a *normalised direction range (NDR) threshold* using the following equation:

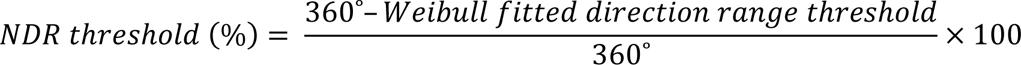

### Home Training protocol

Most participants trained at two, non-overlapping, blind-field locations for at least six months (although a subset – P013, P019 and P020 only trained at one location due to time constraints driven by their particular work/life circumstances). For all training locations, the entire stimulus was within the Humphrey-defined blind field border. NDR thresholds were then measured sequentially, starting with the edge of the stimulus at the perimetrically-defined blind-field border, and moving 1° laterally deeper into the blind field, until performance dropped to chance (∼50% correct) and NDR thresholds became unmeasurable (artificially designated as 100% to represent ceiling performance). Blind field locations selected for training are shown in Figure 2. Participants trained at these locations until performance reached 72% correct, and NDR thresholds stabilised below 100% over 5-10 consecutive sessions. The training location was then moved 1 degree further into the blind field along the x-axis (Cartesian coordinate space).

**Figure 2.**
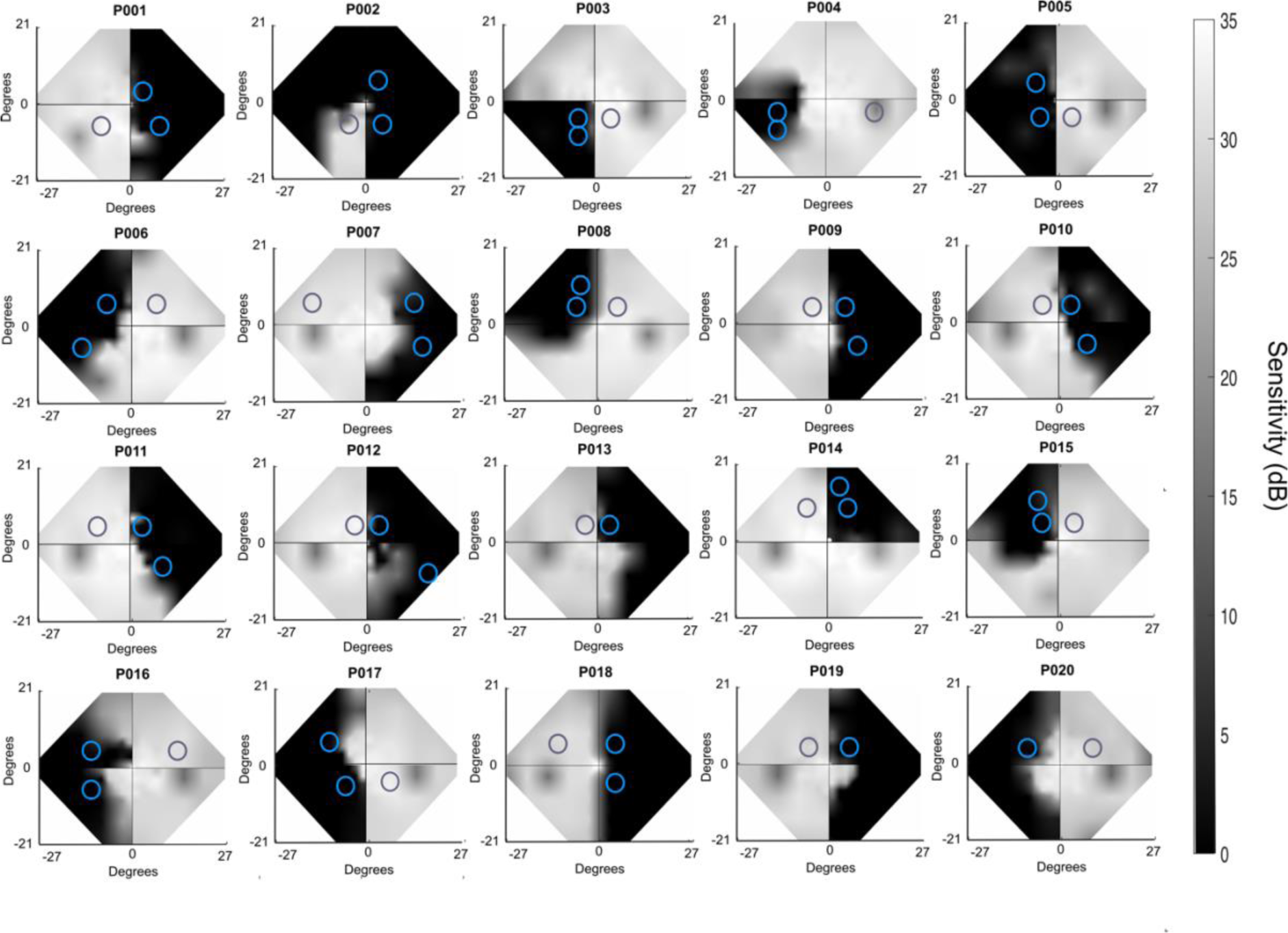
Binocular composite Humphrey Visual Fields. Perimetry from the pre-training visit indicating deficit location. Two blind-field training locations are superimposed on each Humphrey visual field and indicated by blue circles; approximately eccentricity-matched, sighted field locations are denoted by grey circles (three participants only trained at one blind-field location: P013, P019 and P020).

## Behavioural Analysis

### Motion discrimination training

At each research visit, motion discrimination ability on the trained task was measured with fixation enforced using an Eyelink 1000 Plus eye tracker (SR Research Limited, Ontario, Canada). Percentage correct performance and NDR thresholds were calculated for each training location and a matched, sighted-field location. In the majority of cases, when participants trained at 2 blind-field locations for the duration of the study, the location that improved most was used as a measure of improvement capacity in all analyses.

### Humphrey Visual Fields (HVF)

#### Acquisition

A Humphrey Field Analyser was used to collect monocular 24-2 and 10-2 visual fields for each participant at each timepoint. The same two trained researchers (HW and LS) collected all visual field tests. These visual field tests were used to estimate deficit location and severity for each participant. Visual acuity was corrected to 20/20 and eye tracking was controlled to ensure central fixation during this task. Data from participants were excluded if they showed fixation losses, false positives, and negative errors outside of the normal range (≥20%) in either eye at any time point. Data from P001 and P010 were removed from all HVF analyses for these reasons.

#### Pre-processing

As visual field deficits were homonymous, composite binocular HVF were generated by averaging monocular luminance detection thresholds (dB) between eyes, with binocular 24-2 and 10-2 HVFs then combined at overlapping regions (as described previously: (Cavanaugh and Huxlin, 2017; Cavanaugh *et al*., 2019; Willis *et al*., 2024). Luminance thresholds were then interpolated between testing points using a natural-neighbours function to a resolution of 0.1°. Finally, composite HVF were compared between timepoints as follows: pre-training composite fields were subtracted from post-training fields to determine improvements due to training, and post-training composite fields were subtracted from follow-up fields to determine persistence. The area of visual field impairment (deficit size) was defined by points within the blind field with binocular average thresholds below 10dB (the US Social Security Administration definition of impairment; Social Security Administration, 2019). We also calculated the area of the visual field that improved (defined by where detection thresholds in the blind field improved by >6dB: “area improved”) and worsened (defined by where detection thresholds in the blind field worsened by >6dB: “area worsened”). We used the 6dB criterion as this is 2x the test-retest reliability of the HVF (3dB; Cavanaugh and Huxlin, 2017). Finally, maximum and mean luminance detection thresholds were also calculated for each training location based on the interpolated pixels (0.1°) falling fully within the area covered by the training stimulus.

## MRI analysis

### Acquisition

MRI data were acquired on a 3T Siemens Prisma MRI scanner with a 64-channel head coil at the Wellcome Centre for Integrative Neuroimaging, University of Oxford. A structural scan was acquired for each participant at each timepoint. These scans were high resolution whole head T1-weighted anatomical images (1×1×1mm^3^; TE = 3.97ms, TR=1900ms, FoV=192mm, flip angle=8°).

### Anatomical pre-processing

The optic tract volume analysis was adapted from previous studies (Bridge *et al*., 2011; Millington *et al*., 2014; Fahrenthold *et al*., 2021). T1-weighted images were bias-field corrected. FSL software (FMRIB software library) was then used to reorient the optic tracts parallel to the anterior-posterior axis; two coordinates in the optic tract were selected in structural space for each individual scan and a structural to standard space transform was used to reorient the T1 image parallel and perpendicular to the optic tract. Bilateral masks of equal size were hand-drawn over the optic tract in native space slice-by-slice starting posterior to the optic chiasm. Masks were drawn posteriorly until the optic tract was no longer distinct from surrounding structures. The structural image was then masked by these manually-drawn optic tracts to create an ipsilesional and a contralesional optic tract, in which each voxel had a value reflecting the T1-weighted image intensity, with higher values reflecting greater white matter integrity. Three participants were removed from this analysis: P013 due to poor quality MRI images so that accurate OT masks could not be drawn, along with P002 and P010 due to evidence of bilateral damage and therefore no intact, contralesional OT for comparison. P019 had evidence of a potential bilateral stroke in contralesional V2, but since the stroke did not impact V1, they were retained in our analyses.

The volume of each optic tract was then calculated by (1) establishing the maximum voxel intensity across the two optic tracts, and (2) calculating the number of voxels within each mask at 5% intervals between 25 and 95% of this intensity. The laterality index (LI) was then quantified by taking the relative difference in the estimated volume between the two optic tracts - i.e.

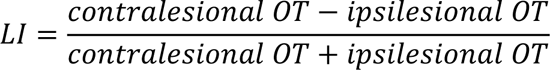

As shown in Figure 3B, the LI for each timepoint was dependent on the threshold used. Once thresholding was increased above 75%, LI gradually increased, since damaged white matter has subtly lower values on a T1-weighted scan. Previous studies used thresholds between 75% (Millington et al., 2014) and 85-95% (Fahrenthold et al., 2021). Millington et al (2014) included patients with long-standing (2-48 years) lesions affecting a range of structures, including the OT, and therefore a threshold of 75% was sufficient to detect differences in white matter. Fahrenthold et al (2021) included stroke survivors with more recent (∼13.4 months post stroke) lesions to V1, and therefore used a threshold of 85-95% to detect smaller differences in white matter in the OT. In the current study, a threshold of 85% was selected for all analyses to optimise the comparison of small differences in white matter integrity of the OT while maintaining the same threshold across participants and timepoints (see in Figure 3A for an example of OT thresholding). To determine the optimum thresholding, we visually inspected the data and took the highest threshold where the structure of the optic tract was maintained. The highest thresholds (such as 95%) resulted in a patchy and disconnected structure, even in the contralesional side, which is more likely to be affected by noise and therefore may not reflect genuine degeneration of the tract (see supplementary Figure S1 for examples).

**Figure 3.**
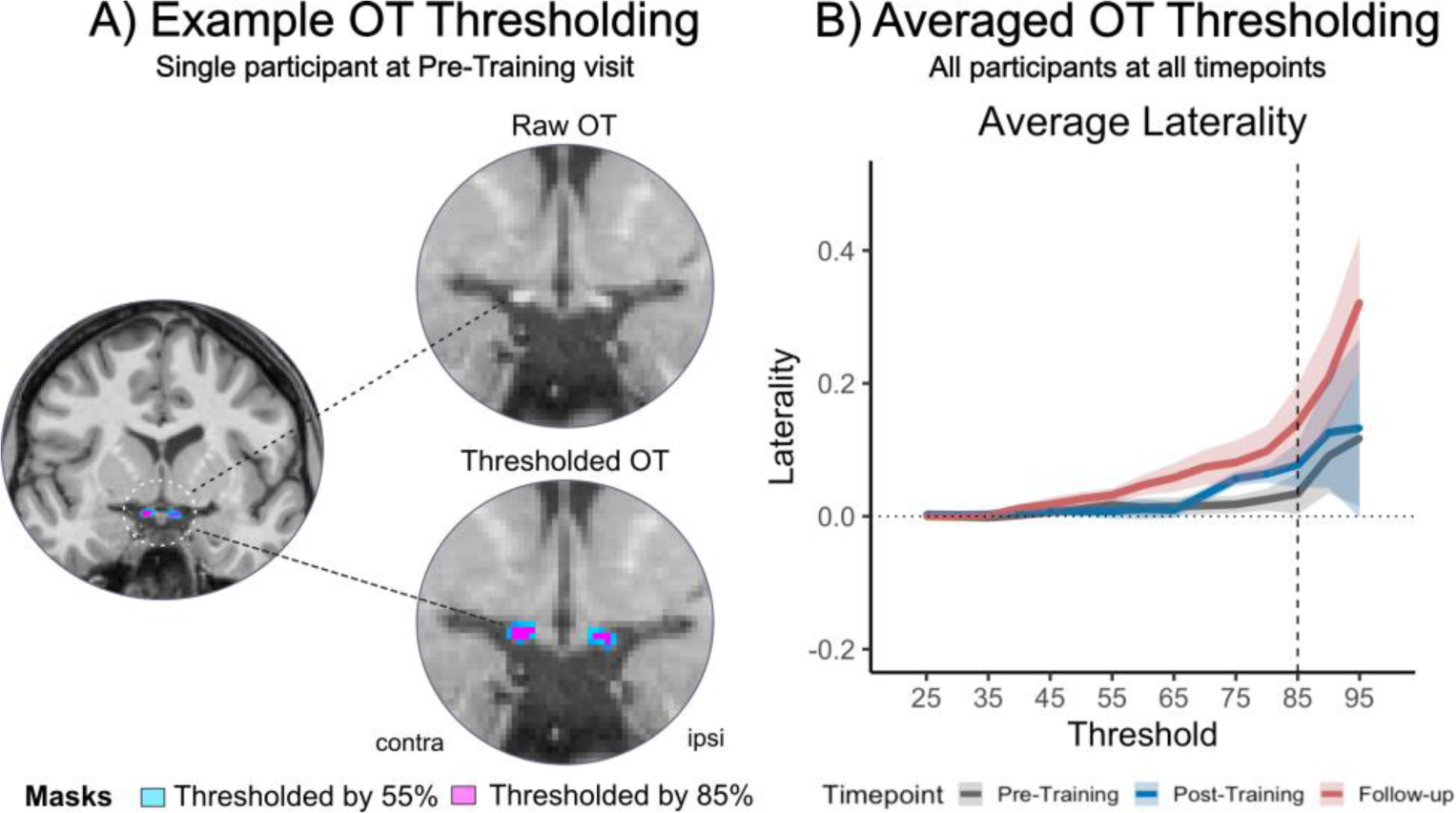
Optic tract structure and thresholding. A) Example of raw OT anatomy and hand-drawn optic tract masks in one representative participant (P001; Pre-Training; contralesional OT on the left; ipsilesional OT located on the right) with thresholding at 55% (light-blue) and 85% intensity (pink). (B) Averaged OT laterality indices over the range of intensity thresholds (25-95%) for the three imaging timepoints (Pre-Training=grey; Post-Training=blue; Follow-up=red). P002, P010 and P013 were removed. Shaded ribbons represent standard error of the mean. Dotted horizontal line indicates a laterality index of 0. Dashed vertical line highlights laterality at 85% threshold.

### Proportion of damage to visual cortex

The proportion of damage to the occipital lobe and ipsilesional V1 was calculated for all participants. Occipital lobe masks were derived from the MNI Structural Atlas in FSL and ipsilesional V1 masks were derived from the extended Human Connectome Project multimodal parcellation atlas (HCPex; (Huang *et al*., 2022). Occipital lobe and V1 masks were registered to individual anatomical space using FNIRT (Andersson, Jenkinson and Smith, 2007). Lesion masks were independently drawn by hand in anatomical space by 2-4 trained researchers (Willis *et al.,* 2023). Researchers included area of damaged tissue within their masks (including damaged grey matter). Lesion definitions were then summed and thresholded so that only voxels identified by at least two researchers were included in the final lesion mask. Occipital lobe and V1 masks were then masked by the lesion using *fslmaths* and the number of voxels within each mask was calculated. The proportion of ipsilesional V1 damage and occipital lobe damage was then calculated for each participant.

### Statistics

All statistical analyses were carried out in R studio (R version 4.1.2) using the *rstatix* package. Data is presented as median *±* interquartile range (IQR) unless otherwise indicated. Where data met the assumptions, parametric tests were used, otherwise non-parametric tests were used. Paired tests (either t-tests or Wilcoxon tests) were used to compare timepoints for the motion discrimination task and OT analysis. Effect sizes are reported for parametric tests (Cohen’s d) and for non-parametric tests (r). For the perimetry and OT analysis, one-sample tests (either t-tests or Wilcoxon tests) were used to compare changes to zero (i.e. no change). P-values were Holm-Bonferroni corrected for multiple comparisons. Participants were removed from analyses if their results differed by more than 3SD from the median.

## Results

### Motion discrimination training: efficacy and persistence

As can be seen in Figure 4, there was limited variability in percentage correct before training (N*=20; median ± IQR* = 53.5*±*4.3%; Figure 4A) or NDR thresholds (Figure 4C; 1*00±0%*) on the motion discrimination and integration task at blind-field locations selected for training. After training, percentage correct performance at these blind-field locations increased significantly (*N=20; 76.5±11.5%;* paired t-test*: t(19)=-7.79, p<0.001; effect size d=-1.74; magnitude=large; Figure 4A*) and NDR thresholds decreased significantly (*N=20; 45.8±58.4%;* paired Wilcoxon*: W=136; p<0.001; effect size r=0.84, magnitude=large*; *Figure 4C*). Four out of 20 participants did not improve on the trained task and their NDR thresholds thus remained unmeasurable at the post-training visit. These results were also reported as behavioural correlates in Willis et al (2024).

**Figure 4.**
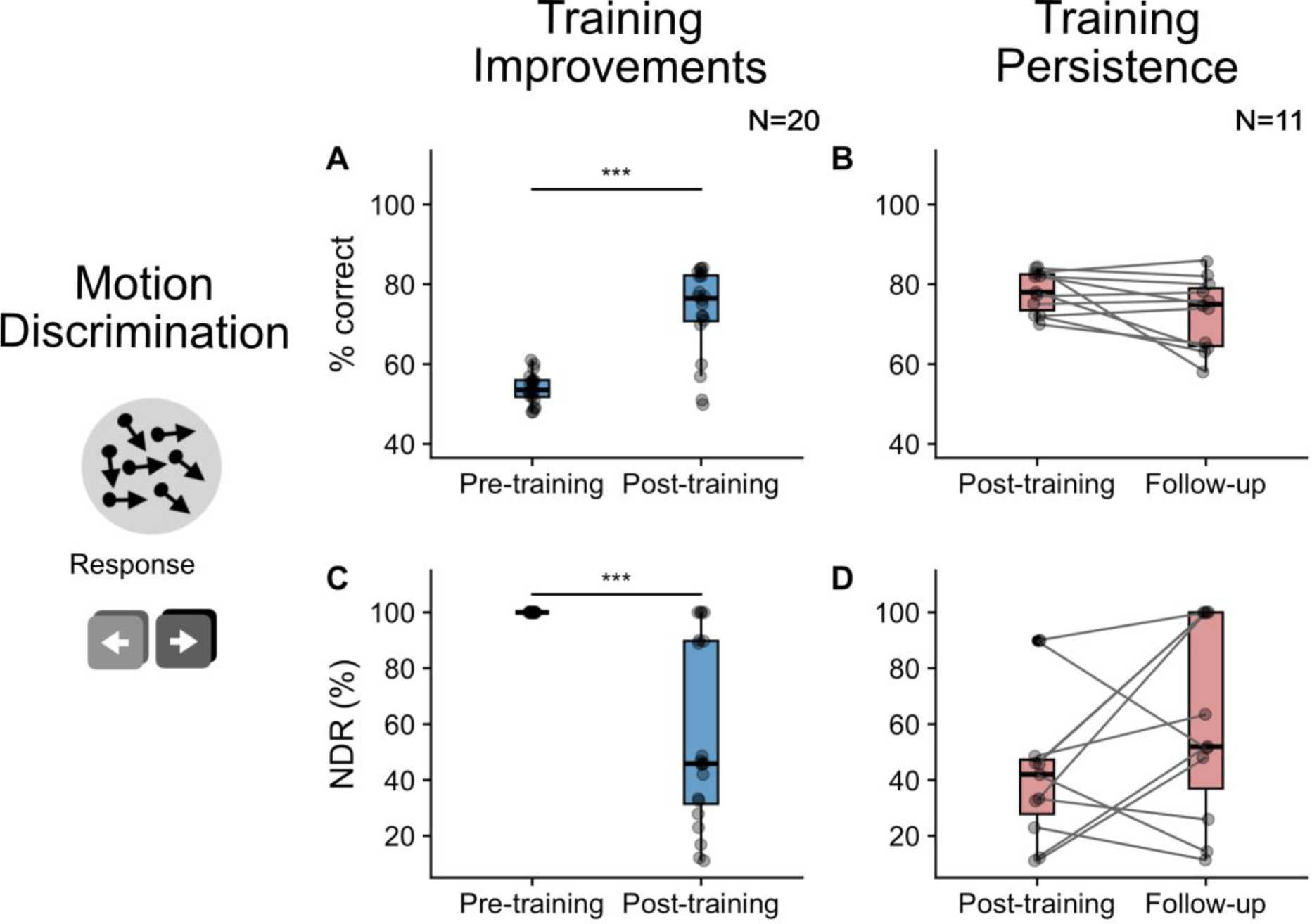
Scatter and box plots indicating recovery and persistence of motion discrimination ability. Change in percentage correct on the task is shown between the pre- and post-training visits (A; blue) and for the period with no training between the post-training visit and follow-up, to quantify persistence of the improvement (B; red). The change in NDR thresholds is also shown between the pre- and post-training visits (C; blue) and post-training and follow-up visits (D; red). Grey scatter points represent the data from each individual participant. Box plots indicate the median (black line) with the upper and lower hinge reflecting the 25% and 75% quantile respectively. Whiskers represent the smallest (lower whisker) and largest (upper whisker) values that are no further than 1.5*IQR from the hinge. Paired tests indicated a significant improvement in both percentage correct and NDR between the pre- and post-training visits (blue), and no significant change (indicating persistence of this effect) between the post-training and follow-up visits (red; significance p<0.001 is indicated with ***). Three participants were removed from persistence plots as they did not improve between the pre- and post-training visits.

We next sought to quantify whether training-induced improvements persisted in the absence of training, at a follow-up visit, conducted ∼3 months later. Three of the participants who failed to improve between pre- and post-training visits were removed from this analysis (their follow-up NDR also remained unchanged at 100%). Percentage correct performance in the remaining 11 participants measured 78*±*9% immediately after training and 75*±*14.5% after a further 3 months without training, generating NDR thresholds of 42.0*±*19.5% and 51.9*±*63.0% respectively (Figures 4B and 4D). While globally there were no significant changes from the post-training visit for percentage correct (paired Wilcoxon*: W=76.5; p=0.06*) or NDR thresholds (paired Wilcoxon*: W=16; p=0.14*), there was clear heterogeneity in outcomes. Specifically, seven out of 11 participants maintained NDR thresholds around 50% - within the range that they attained immediately following training; four participants reverted from measurable NDR thresholds post-training to unmeasurable NDR thresholds (100%) at follow-up.

### Persistence of Humphrey Visual Field changes

Three measures of change in visual field were quantified using Humphrey perimetry: deficit size, area improved, and area worsened. Deficit size was significantly reduced between the pre- and post-training visits *(median ± IQR change=-11.49±22.90* deg^2^*; one-sample t-test: (t(17)=-2.56; p=0.027; effect size r=-0.61, magnitude=moderate; Figure 5A).* Similarly, the area improved between pre- and post-training visits *(13.94±31.01deg^2^)* was significantly greater than zero (*one-sample t-test: t(16)=4.58; p<0.001; effect size r=1.11, magnitude=large; 1 participant removed due to 3SD from mean; Figure 5C).* These results were reported previously as behavioural correlates in Willis et al (2024). Area worsened between the pre- and post-training visits was also significantly different from zero (*3.0±6.8*deg^2^*; one-sample t-test: t(17)=3.49; p=0.002; effect size r=0.82, magnitude=large;* Figure 5E), although the average size of the change was less than the area improved.

**Figure 5.**
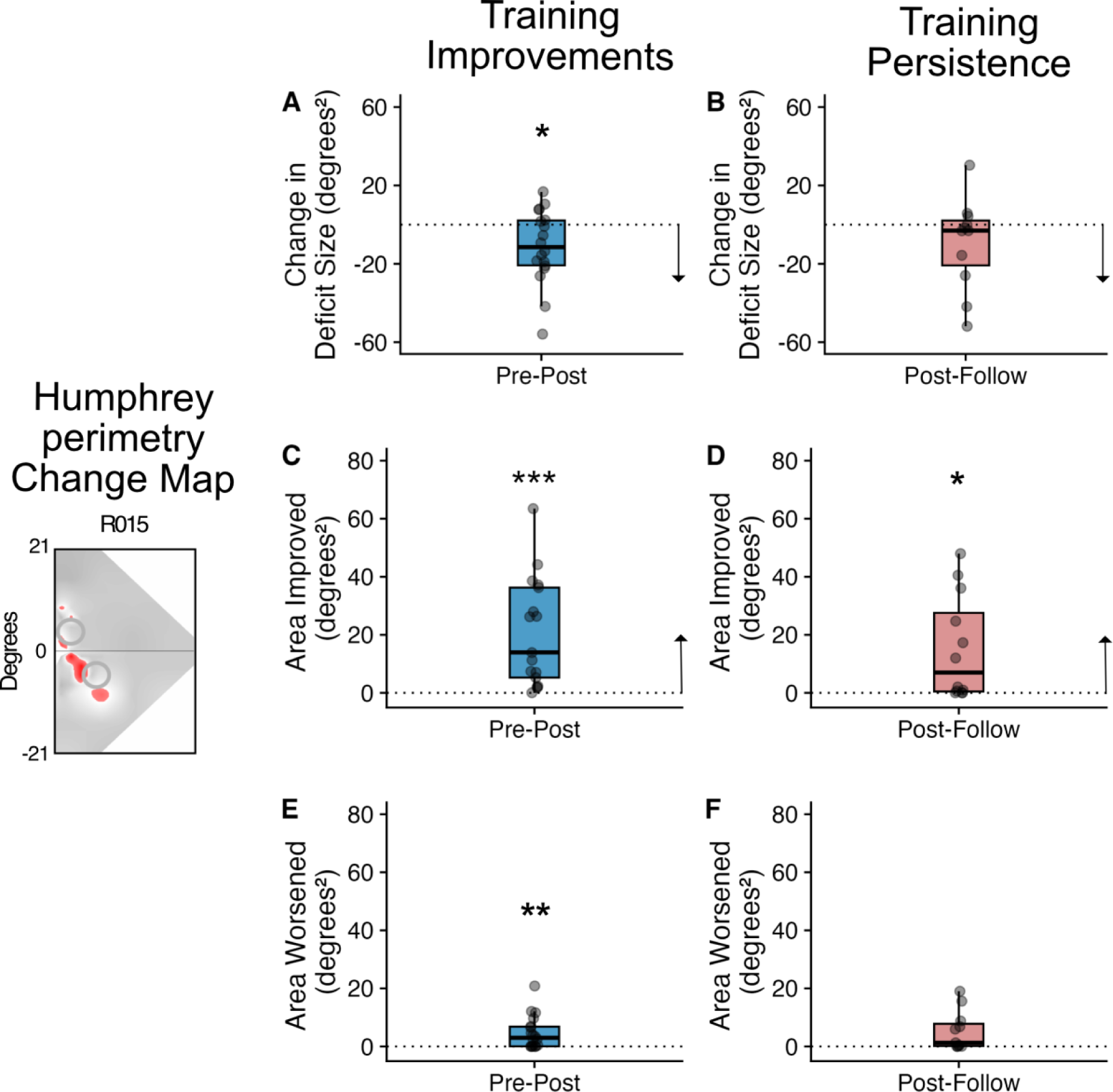
Scatter and box plots indicating change and persistence in Humphrey-perimetry. Humphrey Perimetry Change Map: The left image shows an example of the area of improvement (red) on the perimetry in a single participant (P011). The top row shows the change in deficit size, with A (blue) showing the reduction of the deficit following training. B (red) shows that this reduction in deficit size persists between the end of training and the follow-up scan. C shows the extent of area improved following training, and the persistence of the change measured between the post-training and follow-up session (D). Finally, there were small regions that showed worse thresholds following training (E) which did not change further at the follow-up session (F). Grey points represent the data from each individual participant. Box plots indicate the median (black line) with the upper and lower hinge reflecting the 25% and 75% quantile respectively. The whiskers indicate the smallest (lower whisker) and largest (upper whisker) values that are no further than 1.5*IQR from the hinge. Black arrows indicate directionality of improvements (i.e. lower for change in deficit size, higher for area improved). One-sample tests indicated that there was a significant reduction in deficit size, increase in area of improvement and increase in areas worsened between the pre- and post-training visits compared to zero. There was no significant change in deficit size (indicating persistence of this effect) between the post-training and follow-up visits. Area of improvement showed a slight improvement, while there was no change in area of worsening (Significance indicated by *p<0.05; **p<0.01, ***p<0.001).

While deficit size was significantly reduced between pre- and post-training visits (also reported in Willis et al, 2024), there was no evidence that it changed between the post-training and follow-up visits *(-2.36±22.91 deg^2^; one-sample t-test: t(11)=-0.33; p=0.749; effect size r=-0.09, magnitude=negligible; Figure 5B)*. Moreover, area improved was just significantly greater than zero between post-training and follow-up visits *(7.02±27.08deg^2^; one-sample t-test: t(11)=2.93; p=0.041; effect size r=0.85, magnitude=large; Figure 5D*), suggesting some continued improvements after training had ended. Area worsened between post-training and follow-up visits was not significantly different from zero, showing a slightly lower magnitude of change than the training phase (*1.3±7.63*deg^2^*; one-sample t-test: t(10)=2.59; p=0.054; effect size r=0.78, magnitude=moderate; 1 participant removed due to 3SD from median; Figure 5F).* Thus, globally, it appears that the modest improvements in Humphrey perimetry observed following training also persisted over the 3-months period following cessation of training.

### Optic tract laterality index

Across the three time-points examined (pre-training, post-training, follow-up), the optic tract LI was close to zero (pre-training: 0.03*±*0.18; post-training: 0.06*±*0.13; follow-up: 0.09*±*0.08). LI was not significantly different from zero at the pre-training visit (*N=15; one-sample Wilcoxon test: W=77; p=0.348; effect size r=0.25; magnitude=small; Figure 6A)*. However, by the post-training (*N=15; one-sample Wilcoxon test: W=100; p=0.049; effect size r=0.59; magnitude=large; Figure 6B)* and follow-up visits *(N=10; one-sample Wilcoxon test: W=44; p=0.038; effect size r=0.81; magnitude=large; Figure 6C)* LI became significantly greater than zero, suggesting optic tract degeneration over time across participants. Although both the post-training and follow-up LI measures were significantly greater than zero, a Freidman test showed that there was no consistent change across time points (C^2^(2) = 2.0; p = 0.39).

**Figure 6.**
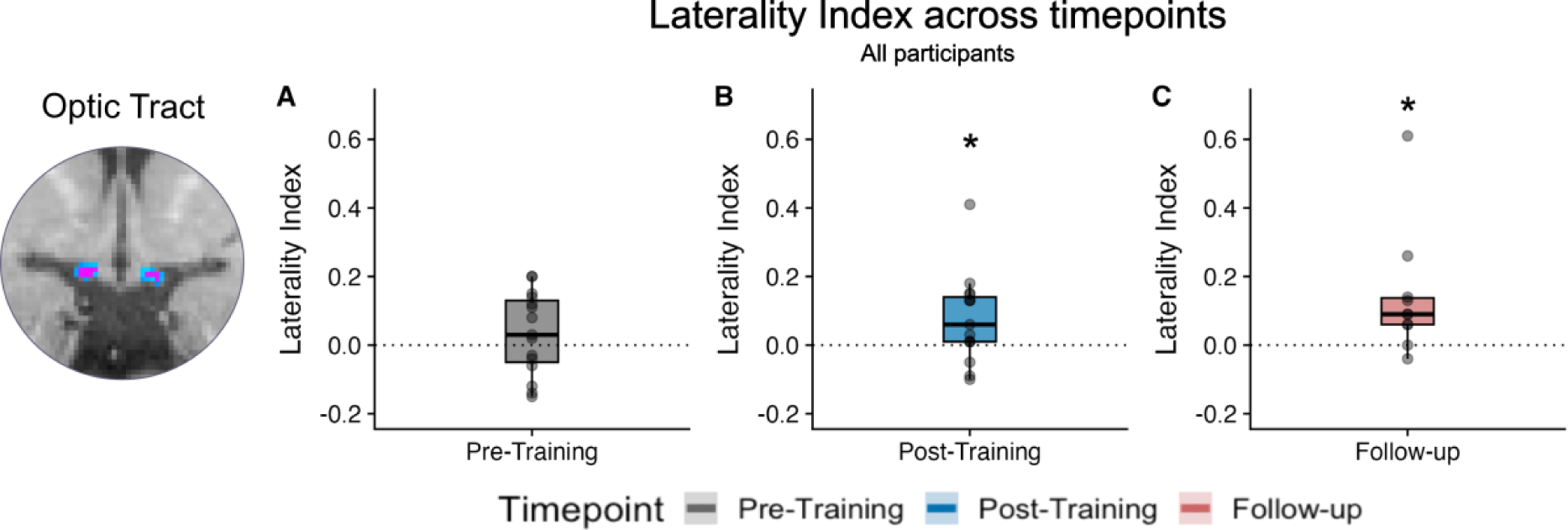
Optic tract laterality indices across timepoints. Laterality indices (85% threshold) for the three timepoints (A: Pre-Training; B: Post-Training; C: Follow-up). Grey scatter points represent the data from each individual participant. Box plots indicate the median (black line) with the upper and lower hinge reflecting the 25% and 75% quantile respectively. The whiskers represent the smallest (lower whisker) and largest (upper whisker) values that are no further than 1.5*IQR from the hinge. One-sample tests indicated that LI values were not significantly different from zero (suggesting limited lateralisation) at the pre-training visit. However, by the post-training and follow-up visits, evidence of increasing lateralisation emerged (indicated by *, p<0.05).

Next, we asked whether lesion size (i.e. proportion of occipital lobe or ipsilesional V1 damage), time since lesion, or deficit size might explain the variance in pre-training LI. As can be seen in Figure 7, linear regression analyses showed no significant relationship between pre-training LI and proportion of occipital lobe damaged (adjusted R^2^=0.08, F[1,13]=2.22; p=0.160; Figure 7A), ipsilesional V1 damaged (adjusted R^2^=-0.1, F[1,13]=0.27; p=0.609; Figure 7B) or deficit size (adjusted R^2^=-0.05, F[1,12]=0.38; p=0.551; Figure 7D). Only time since lesion was strongly, directly related to pre-training LI (adjusted R^2^=0.56, F[1,12]=17.39; p=0.001; Figure 7C). P016 was considered an outlier and removed from this analysis, as their time since lesion was 297 months (all other participants fell within a range of 6–58 months at the pre-training visit). Notably however, LI at the post-training and follow up visits was no longer correlated with time since lesion (post-training: adjusted R^2^=-0.06, F[1,12]=0.21; p=0.657; follow-up: adjusted R^2^=-0.05, F[1,8]=0.55; p=0.48). This suggests that visual training may be disrupting the expected time-related pattern of OT degeneration.

**Figure 7.**
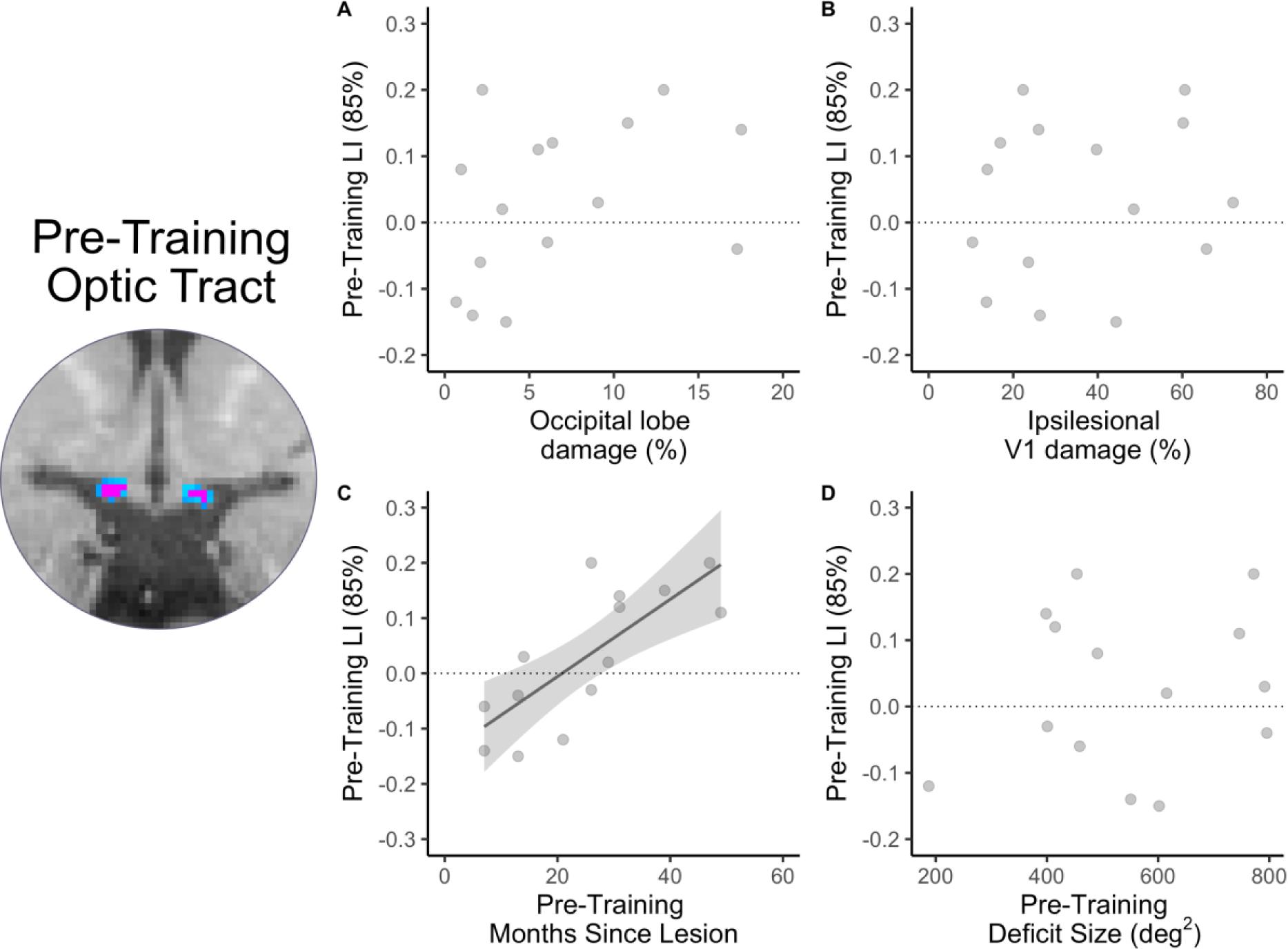
Optic tract laterality indices and covariates. Relationship between pre-training LI and percentage of occipital lobe damage (A), ipsilesional V1 damage (B), pre-training months since lesion (C) and pre-training deficit size (D). There was no relationship between pre-training LI and occipital lobe damage, ipsilesional V1 damage or pre-training deficit size. However, pre-training months since lesion was strongly related to pre-training LI (as indicated by regression line).

Next, we asked whether persistence of learning on the trained task (% correct and NDR) could be explained by a number of covariates. Participants were separated into those who maintained measurable NDR thresholds between the post-training and follow-up visits (“persisted”, N=7), and those who did not (“non-persisted”, N=4). There were only small differences in the median pre-training LI (persisted: 0.14±0.21; non-persisted: 0.06±0.06), follow-up LI (persisted: 0.09±0.06; non-persisted: - 0.07±0.2), participant age in years at the time of stroke (persisted: 49.5±22; non-persisted: 51±19.5), months since lesion at the pre-training visit (persisted: 31±21.5; non-persisted: 28±8) or number of training sessions performed (persisted: 164±17.5; non-persisted: 160±16.5). Those who show persistence of NDR thresholds had on average slightly less damage to V1 (persisted: 26.2±18.8; non-persisted: 57.0±21.8) and smaller deficit sizes (persisted: 505±144; non-persisted: 665±178) than those who did not persist. Although there was some suggestion of differences between groups, the sample size in each group was low, with large variance among participants in all metrics. This suggests that much larger sample sizes are needed to identify factors that determine persistence of training-induced recovery of motion discrimination and integration.

Finally, an important residual question was whether persistence of recovery was related to HVF sensitivity attained at the trained locations. Mean and maximum HVF luminance detection thresholds within the area of the stimulus presented at the analysed training location were calculated for participants with improved NDR thresholds post-training and compared between those with and without persistence at the follow-up visit. There were only small apparent differences between the medians of these groups post-training for both mean (persisted: 2.55±3.19 dB; non-persisted: 3.02±0.48 dB) and maximum HVF luminance detection thresholds (persisted: 10.6±8.11 dB; non-persisted: 12.9±5.74 dB). A similar result was seen at the follow-up visit for both mean (persisted: 2.96±1.58 dB; non-persisted: 4.3±0.37 dB) and maximum thresholds (persisted: 9.11±2.53 dB; non-persisted: 16.5±7.43 dB). Paired tests were not run because of the very limited sample size in each group.

Linear regression analyses were then used to determine whether there was a significant relationship between magnitude of persistence and these covariates. As can be seen in Table 2, there was no significant relationship between persistence of learning on the trained task and pre-training LI, follow-up LI, age, pre-training months since lesion, number of training sessions, proportion of V1 damaged or pre-training deficit size.

**Table 2.**
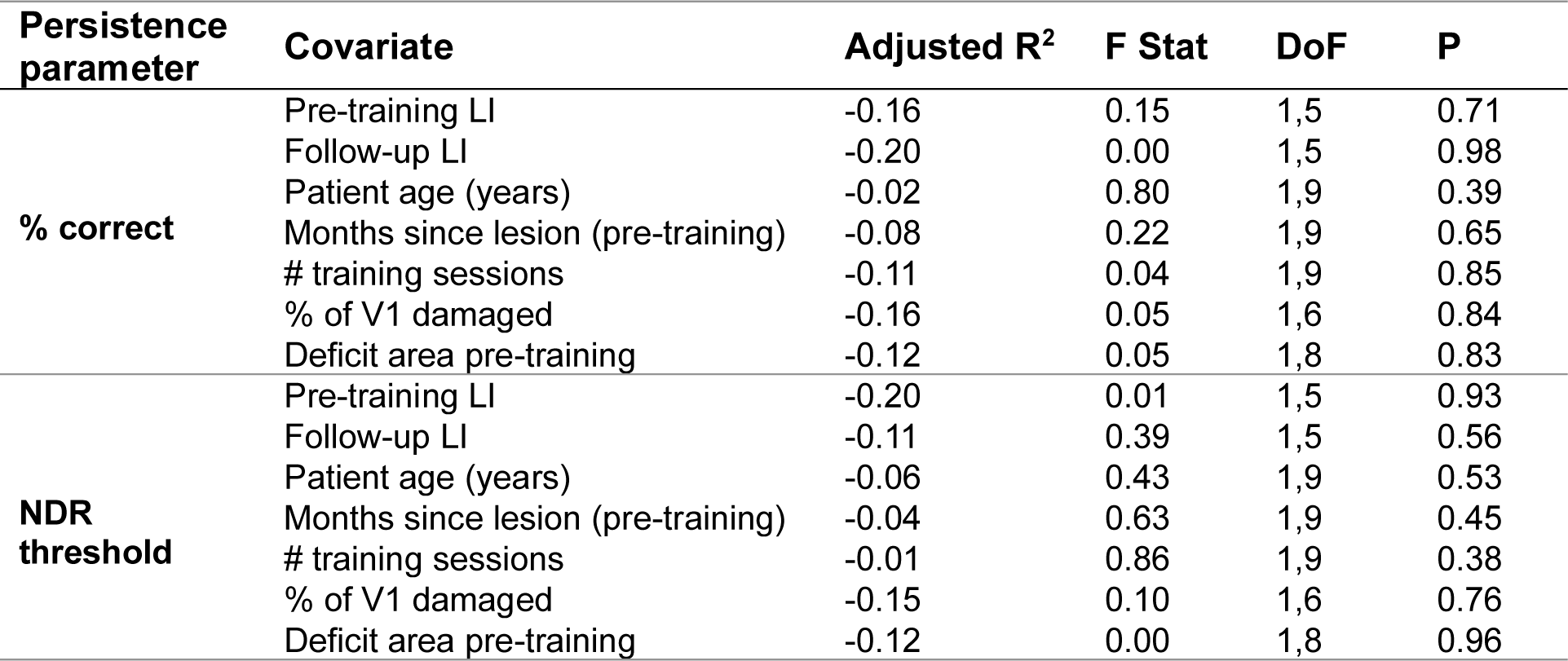
Explaining persistence of trained behaviour. Output of linear regression analyses showing a lack of significant relationship between persistence of improvement in percentage correct and normalised direction range thresholds for the trained task, and covariates (pre-training LI, follow-up LI, age, pre-training months since lesion, number of training sessions, proportion of V1 lesion and pre-training deficit size). NDR: normalized direction range, LI: laterality index, DoF: degrees of freedom.

## Discussion

In stroke survivors with visual field deficits, visual training within the blind field can significantly improve performance on both trained discrimination task and untrained perimetry tasks (Willis et al, 2024). The present study shows, for the first time, that these behavioural improvements persist in most patients after three months without training. However, four patients did not retain their recovered motion discrimination improvements and none of the factors explored (including optic tract degeneration, patient age, stroke age, size of lesion, amount of training or degree of HVF recovery) could explain why they failed to do so.

### Persistence of improvements attained with blind-field training

There is a wealth of research evidence showing that visual training within the blind field after occipital stroke induces perceptual learning that can improve vision for trained tasks (Sahraie *et al*., 2006; Raninen *et al*., 2007; Chokron *et al*., 2008; Huxlin *et al*., 2009; Das, Tadin and Huxlin, 2014; Cavanaugh *et al*., 2015; Joris. Elshout *et al*., 2016; Cavanaugh and Huxlin, 2017; Ajina *et al*., 2021; Willis *et al*., 2024) and generalise to untrained tasks (Sahraie et al., 2006; Chokron et al., 2008; Huxlin et al., 2009; Elshout et al, 2016; Cavanaugh and Huxlin, 2017; Halbertsma et al., 2020; Willis et al, 2024). The present data supports these previous reports, with our training cohort demonstrating significant improvement in both motion discrimination, and luminance detection in the form of perimetry (previously discussed in Willis et al, 2024). However, a key question in the field is whether these improvements persist once training has stopped.

In healthy controls, perceptual learning for motion discrimination tasks has been shown to persist for days (Sundareswaran and Vaina, 1994) and even months after learning (Ball and Sekuler, 1987; Rokem and Silver, 2013; Herpich *et al*., 2019). Here, we report, for the first time, that training-induced recovery of global motion perception appears to persist post-training in the majority of occipital stroke patients. However, four out of eleven of the stroke survivors failed to maintain their initial improvements after three months without training. Even considering our relatively small sample size, this suggests that persistence of visual motion discrimination training may be less robust in occipital stroke survivors than in healthy controls. Alongside recovery of global motion discrimination, improvements in Humphrey perimetry also persisted in our trained cohort after a period without training.

### Persistence of improvement was not related to optic tract integrity or other covariates

Multiple prior studies have provided evidence of OT degeneration in humans and non-human primates with occipital lesions (Bridge *et al*., 2011; Cowey, Alexander and Stoerig, 2011; Millington *et al*., 2014). Previously, LI values between approximately 0 and 1.0 (Millington *et al*., 2014; Fahrenthold *et al*., 2021) were reported in participants with occipital lobe damage, who ranged in time post-stroke from subacute to chronic. In the current study, we saw no overall evidence of OT degeneration at the pre-training visit. However, time since stroke was considerably smaller than in our prior work (e.g. Millington et al., 2014), and a regression analysis within our small sample revealed a strong correlation between degeneration of the ipsilesional OT and the number of months since stroke. The average laterality indices at post-training and follow-up visits also became significantly greater than zero, as would be predicted from the increased passage of time post-stroke.

A prior study found that OT integrity was predictive of rehabilitation success (Fahrenthold et al., 2021). Moreover, ipsilesional retinal ganglion cell complex thickness was reported to be related to early visual cortex activity for blind field stimuli (Schneider et al, 2019). Together, these studies suggest that intact retinal representations of the blind field may be key to supporting visual rehabilitation post-stroke. Here, we asked if retrograde degeneration of early visual pathways impacting the dLGN and then retinal ganglion cells, might limit not only an individual’s capacity for improvement, but also persistence. The limited evidence of OT degeneration pre-training in the current cohort might certainly explain their capacity for improvement, and the persistence of these improvements. However, this is countered by the emergence of OT degeneration at the post-training and follow-up visit, which suggests an ongoing degenerative process. Nonetheless, even at these later visits, LI in the current participants was still much lower (ranging from -0.04 to 0.61 at follow-up) than in the previous studies cited earlier, raising the possibility that visual rehabilitation itself may have had a neuroprotective effect. This is further supported by evidence that at the post-training and follow up visit, LI was no longer correlated with time since lesion, which may suggest that visual training disrupted the natural time course of OT degeneration. Indeed, recent work showed that training within the blind-field of occipital stroke participants appeared to block progressive thinning of the macular ganglion cell and inner plexiform layers (Fahrenthold *et al*., 2024).

Nonetheless, likely related to our small sample size, we found no significant differences in optic tract LI at pre-training and follow-up visits, between participants whose training-induced improvements persisted for three months and those whose improvements did not persist. Although we hypothesised optic tract LI to be the most likely predictor of training persistence after occipital stroke, we also examined the impact of several demographic factors (patient age, time since stroke at training onset) as well as lesion and deficit-specific factors (proportion of V1 damaged, pre-training deficit size), and training-related factors (number of training sessions performed, NDR thresholds attained with training, HVF change and HVF sensitivity attained at trained locations). None of these covariates explained the persistence of training-induced visual improvements or their absence. As with optic tract LI, we speculate that a key contributor for our inability to find significant explanatory factors in the present study was likely our limited sample size relative to the variance across participants. This was in spite of the fact that the present cohort represented a relatively homogeneous sample compared to many prior studies, with participants closely matched in terms of lesion type, time since lesion, lesion size and deficit size (Bridge *et al*., 2011; Cowey, Alexander and Stoerig, 2011; Millington *et al*., 2014). As an aside, homogeneity can also present limitations for exploring variation in OT degeneration, since smaller lesions and deficit sizes tend to be related to limited degeneration (Cowey, Stoerig and Williams, 1999; Cowey, Alexander and Stoerig, 2011; Millington *et al*., 2014). In summary, although this study is the first to explore the persistence of visual rehabilitation in stroke survivors, the limitations imposed by our small sample size mean that future studies with a larger, more diverse sample are needed to answer why some participants maintain training-induced improvements in the blind-field and others do not.

### Limitations

Aside from the small sample size, a number of other issues in the current study also limited our ability to fully understand the neural mechanisms supporting persistence of training-induced improvements in vision post-stroke. For instance, the amount of persistence might have been affected by the magnitude of training-induced improvement. The improvements in the current study were more modest than in previous studies using an identical training paradigm and task, and which reported percentage correct improving to 81% after training (Cavanaugh and Huxlin, 2017), NDR thresholds dropping to 25.5 ± 6.1% (Cavanaugh *et al*., 2019) and HVF areas of improvement around 80 deg^2^. This compares to 76.5±11.5% correct, NDR thresholds of 45.8±58.4% and area of improvement around 13.94deg^2^ in the present study. An important consideration here is that in the above-mentioned previous studies, all participants were allowed to train for as long as needed to improve at a minimum of one location within the blind field (and generally more than one). Here, participants P002, P004, P005 and P019 did not improve on the training task at any location within their allocated timeframe. Given that some participants require many more months to see improvements than others, and it is possible that longer training times (as in previous studies) may have ultimately allowed P002, P004, P005 and P019 to exhibit recovery. A recent clinical trial using a similar training programme and a similar training period (six months) as in the present study reported lower threshold changes (to ∼ 42.1%) and HVF area improved (∼47.9 deg^2^), which more closely approximates the results reported here. Thus, one explanation for the limited persistence in some participants might be that they were not given sufficient training time for improvements to become solidified. Since visual training is highly variable, having strict training schedules might not be the optimum procedure for all patients.

A further explanation for the lack of persistence in some participants might be the selection of sub-optimal training locations. Previous reports have found that some regions of the blind field border are more likely to recover than others (Cavanaugh and Huxlin, 2017; Barbot *et al*., 2021). These studies used extensive mapping of the visual field to select optimal training locations (e.g., Yang et al, 2024). Locations were selected where participants performed slightly above chance performance and were often overlapping with the sighted field. In the current study, we were time limited, so training locations were selected within a few hours, were fully within the blind field border, and were locations where the participant performed at chance. It is possible that in the participants who did not improve and those who did not persist, the locations chosen were less effective (e.g., too deep inside the blind-field). Therefore, further studies are needed to elucidate whether positioning of training locations affects persistence.

### Conclusion

In conclusion, this is the first study to investigate whether improvements after visual rehabilitation in the blind field can persist after a period without training. We found that the majority of participants who significantly improved on the trained task and exhibited reductions in the size of their perimetrically-defined visual field deficit, retained their improvements on both metrics for three months after they stopped training. A small number of participants did not retain improvements in the trained task, but in this small sample, we found no evidence that optic tract degeneration or other demographic, training-specific or deficit-specific covariates could explain this variability in persistence across participants.

## Data Availability

All anonymised data are available on request from the authors following publication.

## Article Information

## Acknowledgments

The authors would like to thank their participants for generously giving up their time to participate. Additionally, the radiographers Jon Campbell, David Parker, Michael Sanders, Nicola Aikin, and Juliet Semple, optometrist Patsy Terry, and optometric technician Charlene Hennesey for assistance with training and data collection.

## Sources of Funding

The project was funded by European Research Council grant to Dr Tamietto (prot. 772953), a British Medical Association Foundation John Moulson Grant and Medical Research Council grant (MR/V034723/1) to H. Bridge. H.E. Willis was supported by the Medical Research Council (MR/N013468/1) and a Waverley Scholarship from The Queen’s College, Oxford. R.S. Millington-Truby is supported by an NIHR academic clinical fellowship. KRH, BKF and MRC were partially supported by a US National Institutes of Health grant (R01 EY027314) co-funded by the Office of Behavioral and Social Sciences Research (OBSSR) and the National Eye Institute (NEI), as well as by an unrestricted grant from Research to Prevent Blindness (RPB) to the Department of Ophthalmology at the University of Rochester. This research was also supported by the NIHR Oxford Health Biomedical Research Centre (NIHR203316). The views expressed are those of the author(s) and not necessarily those of the NIHR or the Department of Health and Social Care. The Wellcome Centre for Integrative Neuroimaging is supported by core funding from Wellcome (203139/Z/16/Z and 203139/A/16/Z). For the purpose of Open Access, the author has applied a CC BY public copyright license to any author-accepted article version arising from this submission.

## Conflicts of Interest

Krystel R. Huxlin is an inventor on US Patent No. 7,549,743. The other authors declare no competing financial interests.

## Supplementary materials

**Figure S1.**
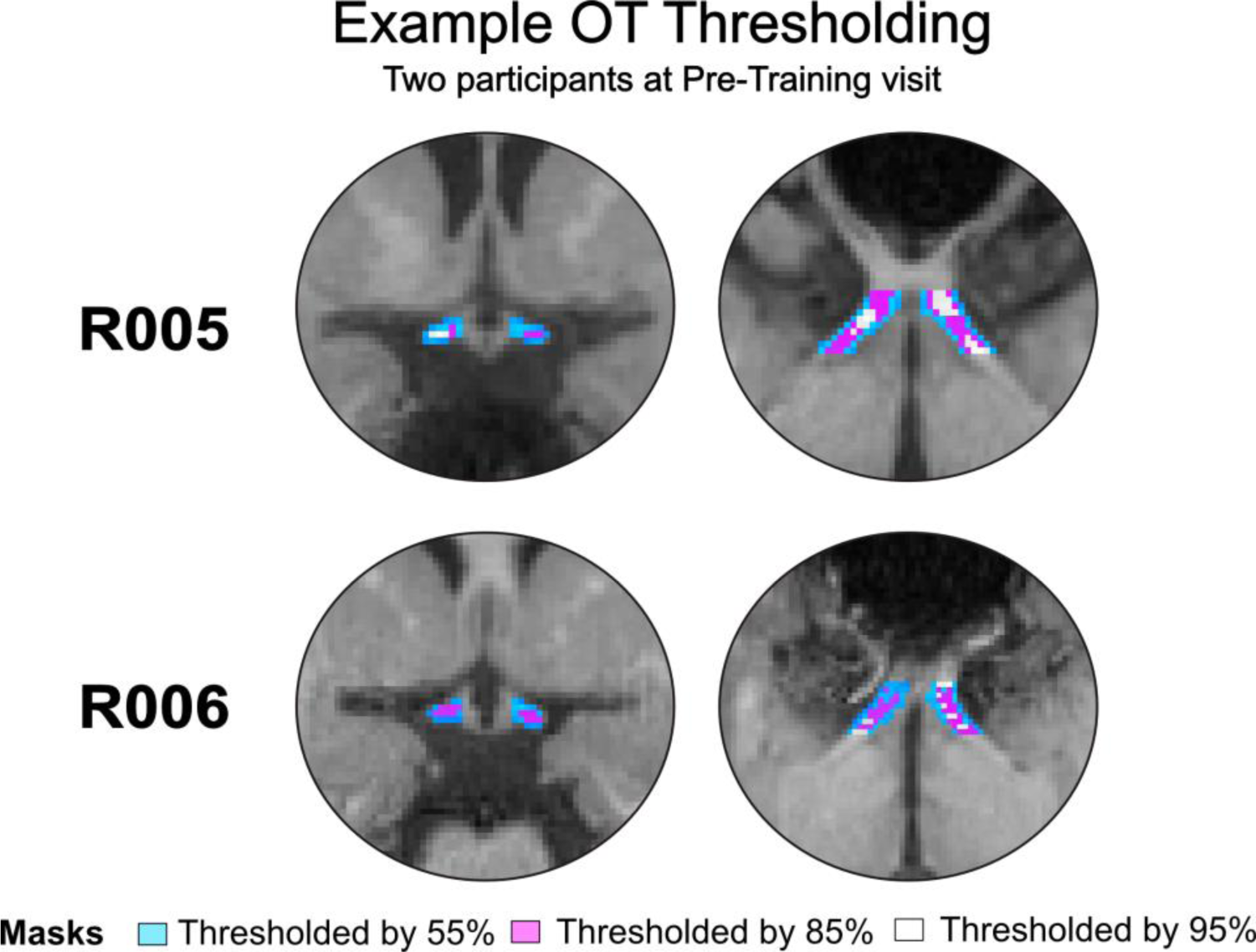
Examples of optic tract structure at higher thresholding. Examples of OT thresholding at 55% (light blue), 85% (pink) and 95% (white). For lower thresholds (55% and 85%) the structure of the optic tract is still maintained. However higher thresholds of 95% results in a patchy and disconnected tract which is likely driven by noise rather than genuine degeneration. For this reason, an 85% threshold was selected.

## Notes

### Clinical Trial

NCT04878861

### Author Declarations

Medical Sciences Interdivisional Research Ethics Committee of University of Oxford gave ethical approval for this work (R60132/RE001).

